# Dengue forecasting and outbreak detection in Brazil using LSTM: integrating human mobility and climate factors

**DOI:** 10.1101/2025.03.02.25323168

**Authors:** Xiang Chen, Paula Moraga

## Abstract

**Background:** Dengue fever is a major global health concern, with Brazil experiencing recurrent and severe outbreaks due to its favorable climate factors, socio-environmental conditions, and increasing human mobility. Accurate forecasting of dengue cases and outbreak risk is essential for early warning systems and effective public health interventions. Traditional forecasting models primarily rely on historical case data and climate variables, often neglecting the role of human movement in virus transmission. This study addresses this gap by incorporating human mobility data into a deep learning-based dengue forecasting framework.

**Method:** An LSTM-based model was developed to forecast weekly dengue cases and detect outbreaks across selected Brazilian cities. The model integrates historical dengue cases, lagged climate variables (temperature and humidity), and human mobility-adjusted imported cases to capture both temporal trends and spatial transmission dynamics. Its performance was evaluated against three alternative models: (1) an LSTM using only dengue case data, (2) an LSTM incorporating climate variables, and (3) an LSTM integrating climate and geographic neighborhood effects. Forecasting accuracy was assessed using Mean Absolute Error (MAE), Mean Absolute Percentage Error (MAPE), and Continuous Ranked Probability Score (CRPS), while outbreak classification was evaluated using accuracy, sensitivity, specificity, and the F1 score.

**Results:** The proposed mobility-enhanced LSTM model consistently outperformed all baselines in both dengue case forecasting and outbreak detection. Across all cities, it achieved lower MAE and MAPE values, indicating improved accuracy, while also demonstrating superior CRPS performance, reflecting well-calibrated uncertainty estimates. In outbreak classification, the model achieved the highest sensitivity and F1 scores, highlighting its effectiveness in detecting outbreak periods compared to models that relied solely on case trends, climate variables, or geographic proximity. The results underscore the importance of integrating mobility data in dengue forecasting, particularly in urban centers with high population movement.

**Conclusion:** By incorporating human mobility dynamics into deep learning-based forecasting, this study presents a scalable and adaptable framework for enhancing dengue early warning systems. The proposed model provides more accurate case predictions and outbreak classifications, offering actionable insights for public health planning and resource allocation. Beyond dengue, this approach can be extended to other vector-borne diseases influenced by mobility and climate factors, supporting more effective epidemic preparedness strategies worldwide.

## 1 Introduction

Dengue fever is a significant global public health threat, particularly in tropical and subtropical regions where environmental conditions favor the proliferation of its primary vector, *Aedes aegypti*. Brazil, with its vast and diverse climate, has consistently reported one of the highest dengue burdens worldwide (Paz-Bailey, Adams, Deen, Anderson, & Katzelnick, 2024; Gurgel-Goņcalves, Oliveira, & Croda, 2024). The disease’s history in the country dates back to the late 19th century, but its modern epidemiology has been shaped by repeated outbreaks and the re-emergence of *Aedes aegypti* after failed eradication efforts. The first major recorded epidemic occurred in Boa Vista, Roraima, in 1981, following the mosquito’s reintroduction in the 1970s (Figueredo, 2003; Braga & Valle, 2007). Since then, dengue has become endemic across the country, with recurring seasonal epidemics affecting millions of people.

The year 2024 marked an unprecedented dengue outbreak in Brazil, with approximately 6.6 million probable cases and 6,199 deaths reported by December 2024 (Brazil, 2025). This surge in cases not only underscored the increasing public health burden of dengue but also highlighted its geographical expansion into previously unaffected areas. Notably, southern municipalities, which historically reported lower dengue incidence due to cooler climates, experienced substantial case increases, demonstrating the disease’s evolving transmission dynamics (Souza et al., 2024). A combination of climatic, demographic, and socio-environmental factors facilitates the persistence and spread of dengue in Brazil. The country’s tropical and subtropical climate provides an ideal environment for mosquito breeding, while rapid urbanization, inadequate infrastructure, and population mobility further exacerbate the risk of outbreaks.

Given the escalating dengue burden in Brazil, forecasting models play a crucial role in guiding public health responses. Traditional dengue prediction methods often rely on epidemiological and climate-based models, leveraging variables such as temperature, precipitation, and humidity to anticipate outbreak patterns. Dengue forecasting has evolved through the application of diverse methodologies, ranging from statistical time-series models to advanced machine learning techniques. Classical models such as ARIMA and SARIMA have been widely used to capture trends and seasonality in dengue incidence (Luz et al., 2008; Cortes et al., 2018; Silawan et al., 2008; Buczak et al., 2018). While effective in stable transmission settings, these models struggle with non-linear relationships, abrupt outbreak surges, and external factors such as climatic shifts and socio-environmental changes. To address these limitations, machine learning approaches—including LSTMs and other deep learning architectures—have emerged as promising alternatives, leveraging their ability to learn complex temporal dependencies and improve predictive accuracy (Chen & Moraga, 2025a; Roster, Connaughton, & Rodrigues, 2022; Kakarla et al., 2023; Zhao et al., 2020; Majeed, Shafri, Zulkafli, & Wayayok, 2023). However, many of these studies primarily focus on epidemiological time series and climate variables, often neglecting spatial dependencies and human mobility, which are critical for understanding dengue transmission beyond local patterns. The inclusion of climate data has significantly improved dengue forecasting accuracy, as meteorological factors directly influence the *Aedes aegypti* mosquito lifecycle. Temperature, humidity, and precipitation impact mosquito breeding, survival, and virus transmission capacity, making them essential predictors of dengue outbreaks. Both statistical and machine learning models have demonstrated improved performance when integrating climate variables, outperforming purely case-based models in multiple regions, including Brazil, Colombia, Mexico, and Myanmar (Johansson, Reich, Hota, Brownstein, & Santillana, 2016; Ortega-Lenis, Arango-Londoño, Hernández, & Mor-aga, 2024; Pavani, Bastos, & Moraga, 2023; Zaw et al., 2023). The delayed effects of climate on dengue cases have also been well-documented, with studies identifying optimal lag structures that capture the time required for environmental conditions to affect mosquito dynamics and disease transmission. While these advancements have strengthened predictive capabilities, climate-driven models still operate under the assumption of localized transmission and fail to account for the role of human mobility in spreading dengue beyond environmentally favorable areas.

To address spatial dependencies, previous studies have incorporated case data from neighboring regions into forecasting models, recognizing that dengue transmission is not confined to administrative boundaries (Moraga, 2019). Spatial models such as geostatistical approaches and spatial autoregressive models have demonstrated improved accuracy by capturing regional correlations and disease spread patterns (Thiruchelvam, Dass, Asirvadam, Daud, & Gill, 2021; Cuong et al., 2013; Moraga, 2023; Chen & Mor-aga, 2025b). However, these methods largely rely on static spatial relationships, assuming that geographical proximity alone dictates transmission dynamics. In reality, dengue spread is heavily influenced by human movement, as infected individuals can introduce the virus into new areas where competent mosquito vectors are present. Traditional spatial models do not explicitly account for this mobility-driven transmission, limiting their ability to predict outbreaks in regions that are not immediate neighbors but are strongly connected through human travel.

This gap underscores the need for forecasting models that incorporate human mobility data to better capture the true dynamics of dengue transmission. While some studies have begun integrating mobility data—such as airline travel and commuting patterns—into infectious disease modeling (Moraga et al., 2019; Mahmood, Amaral, Mateu, & Moraga, 2022; Oliveira et al., 2024), their application in dengue forecasting remains limited (Poongavanan et al., 2024; Kraemer et al., 2019; Kiang et al., 2021; Bomfim et al., 2020; Islam & Hu, 2024). Most existing models still rely on either climate-adjusted epidemiological trends or static spatial effects, overlooking the influence of human travel on dengue propagation. By addressing this limitation, forecasting models can provide more realistic, actionable insights for public health interventions.

This study aims to enhance dengue forecasting and outbreak detection in Brazil by integrating climate factors and human mobility data into a predictive modeling framework. Although the LSTM model itself is well established, our contribution lies in designing epidemiologically meaningful inputs — reflecting both temporal and spatial transmission drivers — and demonstrating their value for dengue forecasting. While traditional models primarily rely on historical case data and climate variables, they often fail to account for mobility-driven transmission dynamics, which play a crucial role in introducing and sustaining outbreaks across regions. By incorporating human movement data alongside climate information, we aim to develop a more robust predictive model capable of capturing both temporal trends and spatial transmission pathways. This integration not only refines outbreak predictions but also provides actionable insights for targeted public health interventions, ultimately supporting more effective dengue control strategies.

In this study, we propose an enhanced Long Short-Term Memory (LSTM) neural network model to predict dengue cases at the city level across Brazil. The model integrates a diverse set of lagged climate variables, including temperature and humidity, to account for environmental influences on mosquito dynamics and viral transmission. Additionally, we incorporate human mobility-adjusted imported dengue cases, allowing the model to capture cross-regional transmission effects driven by human movement patterns (Figure 1). This design allows the model to better reflect real-world transmission pathways and environmental conditions.

**Figure 1:**
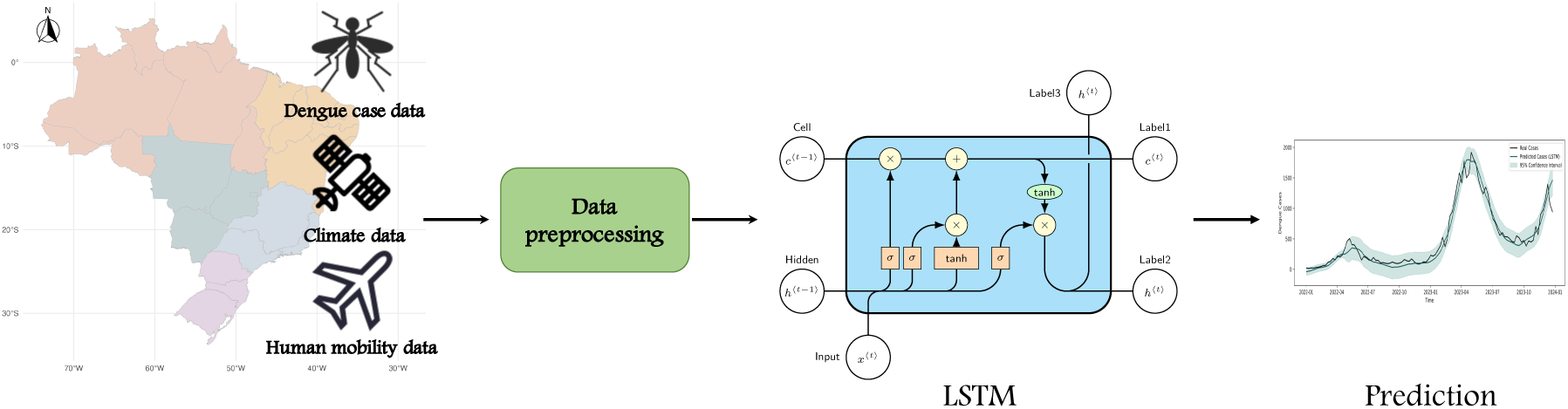
Data processing pipeline for the forecasting of dengue cases in Brazil.

To evaluate the model’s performance, we compare it against three alternative models, including simpler LSTM-based approaches, across two key tasks: forecasting dengue case counts and detecting outbreak periods. For the first task, predicting dengue case counts, model performance is assessed using Mean Absolute Error (MAE), Mean Absolute Percentage Error (MAPE), and Continuous Ranked Probability Score (CRPS) to comprehensively evaluate accuracy and uncertainty in the forecasts. The MAE and MAPE measure absolute and relative prediction errors, while CRPS assesses the calibration and sharpness of probabilistic forecasts, ensuring that prediction intervals are well-aligned with actual dengue cases.

For the second task, detecting dengue outbreaks, we compare the models’ ability to detect outbreak periods using accuracy, sensitivity, specificity, and the F1 score. These metrics provide a balanced evaluation of the models’ ability to correctly classify both outbreak and non-outbreak weeks, minimizing false positives while ensuring critical outbreaks are detected. By evaluating both numerical forecasting accuracy and outbreak classification performance, we assess the added value of human mobility data, demonstrating its role in improving predictive precision, refining outbreak detection, and enhancing early warning capabilities for dengue control.

## 2 Methodology

### 2.1 Selected Brazilian cities

Brazil is a vast country with diverse climatic zones and epidemiological patterns that influence the transmission of dengue fever. To ensure a comprehensive and representative study, we selected 10 major cities across different regions of Brazil, considering historical dengue burden, climate variability, urbanization, and recent outbreak trends. These cities encompass areas with traditionally high dengue incidence as well as locations experiencing a southward expansion of dengue cases. By including cities from different climatic zones and epidemiological profiles, the study accounts for the diverse transmission patterns observed across the country, enhancing the robustness and generalizability of the forecasting models.

Figure 2 presents the geographical distribution of the 10 selected cities for dengue forecasting across Brazil. The selected cities and their respective states are Manaus (Amazonas - AM), Beĺem (Pará - PA), Fortaleza (Ceará - CE), Salvador (Bahia - BA), Braśılia (Federal District - DF), Goîania (Goiás - GO), Belo Horizonte (Minas Gerais - MG), Rio de Janeiro (Rio de Janeiro - RJ), Sāo Paulo (São Paulo - SP), and Curitiba (Paraná - PR). These cities represent a diverse range of climatic conditions, urban environments, and disease transmission dynamics as follows:

- Northern Region (Manaus, Beĺem): Characterized by an equatorial climate, high humidity, and persistent rainfall, which provide a favorable environment for *Aedes aegypti* proliferation.
- Northeastern Region (Fortaleza, Salvador): Experiences tropical and semi-arid climates, with seasonal dengue outbreaks driven by fluctuations in rainfall and temperature.
- Central-West Region (Braśılia, Goiânia): Features a tropical savanna climate, with a distinct dry season and strong temperature variations, influencing mosquito breeding and disease transmission.
- Southeastern Region (Belo Horizonte, Rio de Janeiro, São Paulo): Highly urbanized with humid subtropical and tropical climates, making it a significant contributor to national dengue cases.
- Southern Region (Curitiba): Traditionally less affected by dengue, but recent epidemiological shifts indicate an increasing number of cases, likely driven by climate change and urbanization.

**Figure 2:**
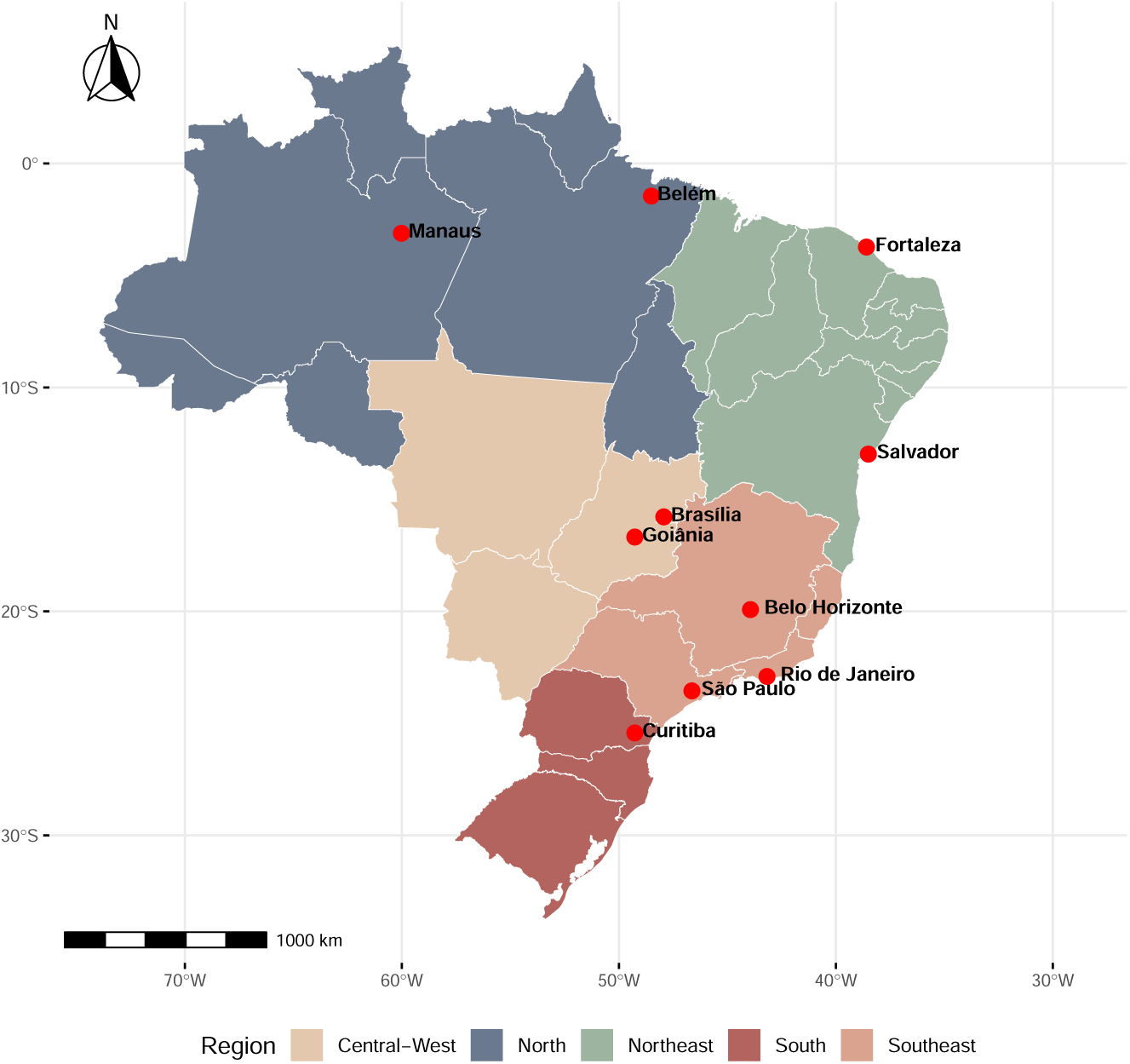
Geographical distribution of selected 10 cities for dengue forecasting

In summary, the selected 10 cities represent a deliberate and strategic sample that balances geographic diversity, epidemiological relevance, and computational feasibility. This enables the evaluation of our framework across distinct climatic zones, transmission dynamics, and urban contexts. While we limit the scope to these 10 locations for tractability, the underlying methodology is highly generalizable. As discussed later, our framework can be extended to other cities, regions, and even countries, wherever comparable mobility, climate, and epidemiological data are available. This design ensures both methodological rigor and practical scalability for future applications.

### 2.2 Data

Dengue data are obtained from InfoDengue (Codeco et al., 2018), a surveillance system that compiles dengue case reports from official health sources across Brazil. This dataset provides weekly dengue cases at the municipal level. InfoDengue also supplies climate data, including temperature and humidity, which are key environmental factors influencing mosquito population dynamics and dengue transmission. Additionally, population estimates from InfoDengue are used to normalize dengue cases and compute mobility-adjusted case importation.

Table 1 provides a summary of the average weekly dengue cases and climate data from 2016 to 2023, along with population characteristics from the 2022 census for the ten selected cities. Figure 3 shows the temporal trends of weekly dengue cases from 2016 to 2023 for the ten selected cities, highlighting seasonal variations and potential outbreak periods.

**Figure 3:**
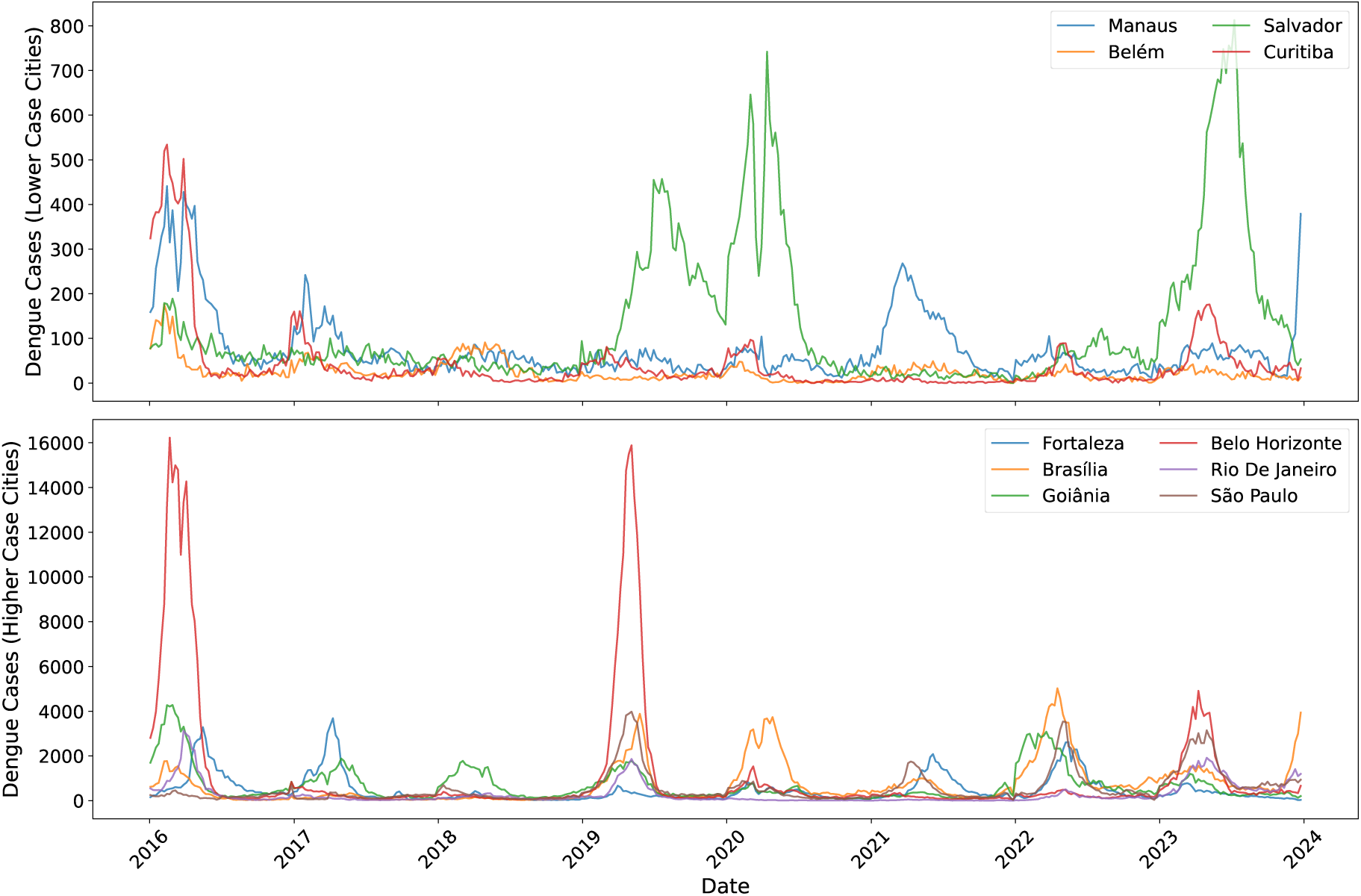
Weekly dengue cases for selected cities (2016-2023).

**Table 1:**
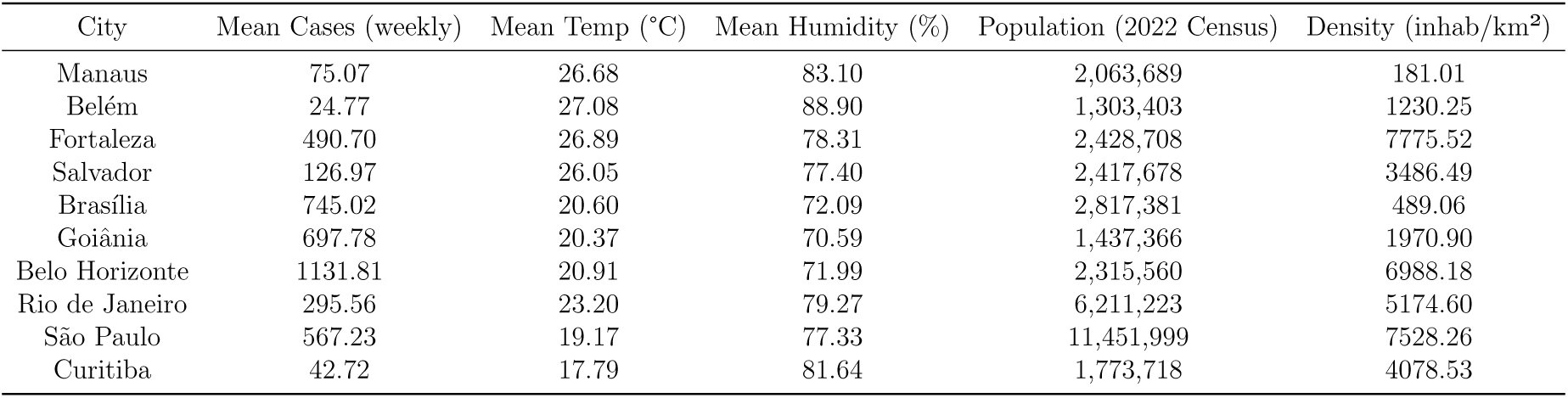
Summary of weekly dengue cases, climate, and population data from 2016 to 2023.

Human mobility data are sourced from (Oliveira et al., 2024), which integrates multimodal transportation data, including air, water, and road travel. This dataset captures intercity movement patterns, enabling the estimation of dengue importation through human travel. To construct the mobility dataset, large-scale intercity travel data were compiled from multiple administrative sources. Road and river transport data (2016) were obtained from the Brazilian Institute of Geography and Statistics, intercity bus capacity data (2022) were sourced from the Brazilian National Transport Confederation’s 2022 transport report, and air travel data (2017–2023) were collected from the National Civil Aviation Agency. Using this information, a graph-based representation of Brazil’s mobility network was developed, where nodes represent cities and edges represent the intensity of human movement across different transportation modes. By incorporating these mobility flows into the forecasting model, we aim to enhance predictions by accounting for the spatial spread of cases driven by human movement.

Figure 4 presents an overview of human mobility patterns in Brazil, where the intensity of connections reflects the strength of mobility flows between cities. This visualization underscores the role of travel in dengue transmission and the importance of integrating mobility data into forecasting models. Mobility is highly concentrated in major urban centers, such as Sāo Paulo, Rio de Janeiro, Braśılia, and Salvador, which serve as key transit hubs facilitating large-scale human movement. These high-mobility regions are particularly relevant for dengue transmission, as frequent travel between cities can introduce infected individuals into new areas, sustaining or amplifying out-breaks.

**Figure 4:**
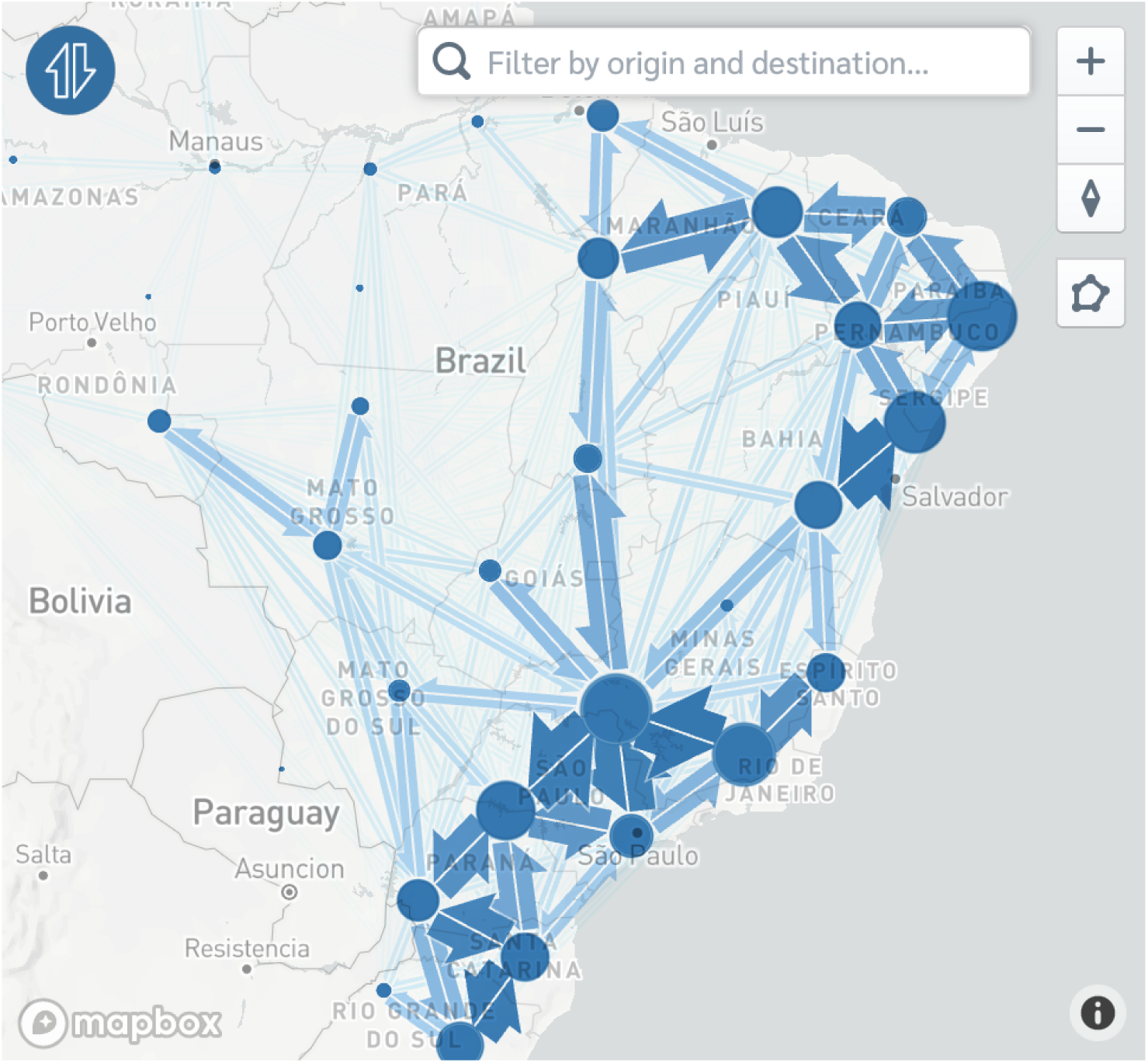
Human mobility patterns describing intercity connection.

### 2.3 Study design

We develop an LSTM-based model to forecast dengue cases and detect outbreaks in Brazil by integrating historical dengue case data, climate variables, and human mobility data. The LSTM model architecture is designed to capture long-term temporal dependencies while integrating spatial transmission signals through mobility data. Unlike previous approaches that primarily relied on spatial effects from neighboring regions (Chen & Moraga, 2025b; Bomfim et al., 2020), our model explicitly incorporates mobility-driven transmission dynamics by quantifying the contribution of dengue cases imported into a given city. This enhancement allows for a more comprehensive representation of dengue transmission patterns influenced by population movement.

#### 2.3.1 Forecasting

The study focuses on medium-term forecasting horizons, generating predictions and uncertainty intervals for one month (four weeks) ahead. Shorter-term forecasts, such as one-week predictions, may lack the necessary lead time for effective public health responses, whereas monthly forecasts provide actionable insights, enabling authorities to implement timely interventions.

To train the dengue prediction model, we employ a moving window strategy with a fixed window size of seven years. The initial training period spans from January 3, 2016 (the first epidemiological week of 2016), to December 25, 2022 (the last epidemiological week of 2022), comprising 364 weeks (52 weeks per year × 7 years). The window is then moved forward in four-week increments to iteratively generate forecasts for 2023 (Figure 5). The model generates multi-horizon forecasts for the next four weeks at each prediction step. The input window is then advanced in 4-week increments to iteratively produce monthly predictions for 2023, ensuring consistency between prediction horizon and evaluation intervals. We chose a fixed-length window to ensure consistent input dimensions, reduce model complexity, and focus learning on recent epidemiological patterns. While including more distant historical data might capture rare large outbreaks, it could also introduce outdated transmission dynamics due to changes in immunity levels, vector behavior, and public health interventions. This strategy balances model simplicity with adaptability to evolving trends.

**Figure 5:**
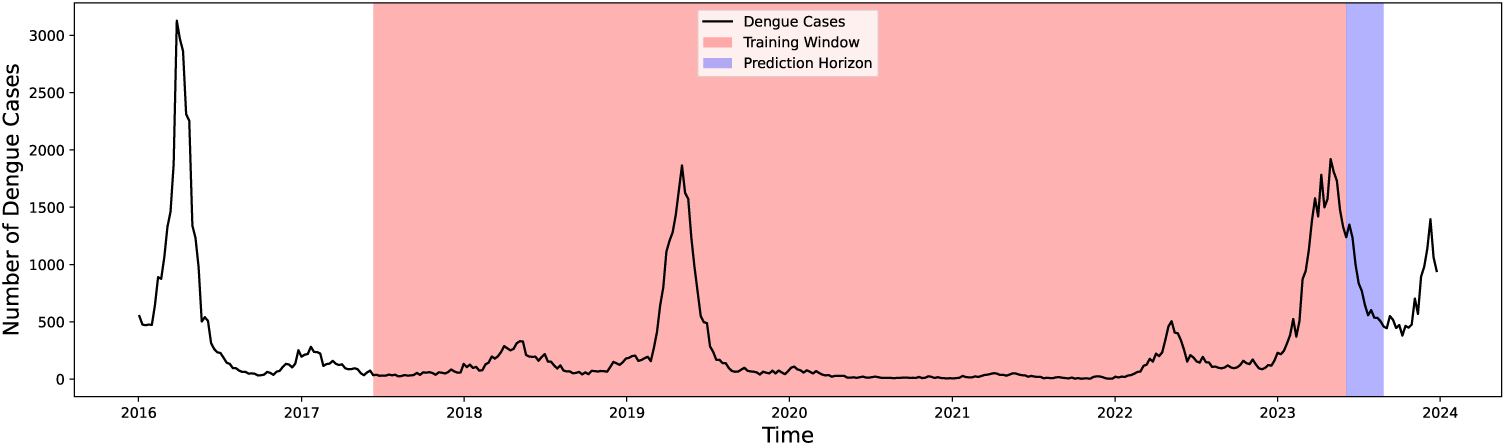
Illustration of the moving window strategy for dengue case forecasting.

#### 2.3.2 Detection of outbreaks

Reliable outbreak prediction is essential for early warning systems, enabling public health authorities to allocate resources and implement targeted interventions effectively. In this study, we not only forecast dengue case counts but also detect dengue outbreaks. The same LSTM model is used for both tasks, with its predicted weekly case counts serving as the basis for outbreak detection.

To determine whether a given epidemiological week in 2023 constitutes an outbreak, we apply two threshold-based strategies derived from historical dengue data. An outbreak is declared if either the observed number of dengue cases or the lower bound of the model’s predictive interval exceeds the corresponding threshold, ensuring that even the most conservative forecast indicates an elevated risk. The first outbreak threshold is based on the mean historical dengue cases over the past seven years (from 2016 to 2022). Thus, for each epidemiological week *i*, the outbreak threshold is calculated as:

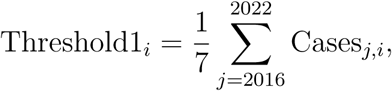

where Cases*_j,i_* represents the reported dengue cases in epidemiological week *i* of year *j*.

While this method provides a baseline measure, it may underestimate outbreaks in years with increasing dengue trends or overestimate them if past outbreaks caused extreme values in the dataset. To account for variations in historical data, we introduce a second, more adaptive threshold that incorporates both the mean and variability in historical dengue cases. For each epidemiological week *i* in 2023, the threshold is computed as:

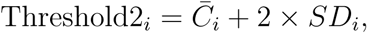

where *C_i_* is the mean number of dengue cases reported during epidemiological week *i* across the years 2016–2022 (i.e. the previous Threshold1 above); and *SD_i_* is the standard deviation of dengue cases for that epidemiological week over the same period. This approach accounts for natural variability in dengue transmission, making it more sensitive to significant deviations from normal seasonal trends. By adopting two threshold definitions—a simple historical mean and a more adaptive threshold accounting for interannual variability—and integrating uncertainty-aware predictions, this framework enables both operational relevance and statistical robustness. This dual approach enhances outbreak detection and early warning capabilities, supporting more proactive and context-sensitive public health responses.

### 2.4 LSTM forecasting model

The LSTM model proposed incorporates three primary sources of information. Namely, (1) historical dengue cases, providing temporal trends of weekly dengue cases for each city; (2) lagged climate variables including temperature and humidity variables at selected time lags, capturing climatic influences on mosquito dynamics and disease transmission; and (3) imported dengue cases, representing the external infection pressure from connected cities.

#### 2.4.1 Incorporating human mobility into the forecasting model

Human mobility plays a crucial role in dengue transmission, as infected individuals can introduce the virus into new regions where competent mosquito vectors are present. Unlike traditional forecasting models that primarily rely on historical dengue cases and climate variables, our modeling approach explicitly accounts for imported dengue cases due to human mobility, enhancing predictive accuracy.

Specifically, to quantify mobility-driven disease spread, the model incorporates an estimate of the imported dengue cases for each city *i* in epidemiological week *t* = 1, 2*, …,* 52 in 2023 that is obtained as follows:

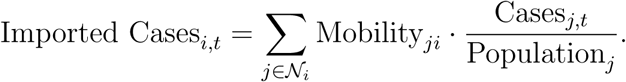

Here, Cases*_i,t_* and Cases*_j,t_* are the number of dengue cases at week *t* in cities *i* and *j*, respectively, and Population*_j_* is the population of city *j*. N*_i_* represents the set of cities *j* that have mobility connections with city *i*; and Mobility*_ji_* is the mobility flow from city *j* to city *i*.

This formulation adjusts the incoming dengue burden based on the prevalence of dengue at the origin city and the relative mobility flow, allowing for a more realistic representation of imported cases. By incorporating this measure, the model dynamically accounts for inter-city transmission risks, which are often overlooked in traditional spatial models that rely solely on geographic proximity. While we do not claim causal inference, prior studies have shown that mobility patterns are empirically associated with dengue spread (Islam & Hu, 2024), and our goal is to leverage these associations to improve predictive accuracy within a practical forecasting framework.

#### 2.4.2 Forecasting process

Figure 1 outlines the entire forecasting process. The process begins with the collection of dengue case, climate, and mobility data, followed by their integration into a structured dataset. In the data preprocessing stage, climate variables are lagged to account for delayed effects on mosquito populations, and mobility-adjusted dengue case importation is computed. Once preprocessed, the dataset is formatted into sequential input windows suitable for the LSTM model, where each input instance consists of the past seven years of dengue cases, lagged climate variables, and imported cases outside, with the model predicting dengue cases four weeks ahead.

As depicted in Figure 6, the LSTM model consists of the following components. First, the input layer receives the sequential input of dengue cases, climate variables, and imported cases. Then, LSTM layers process the sequential data, leveraging gated memory cells to retain long-range dependencies and filter out irrelevant information. The dense layers extract higher-level features and map them to the final output. Finally, the output layer produces the predicted dengue cases for the target city, four weeks ahead.

**Figure 6:**
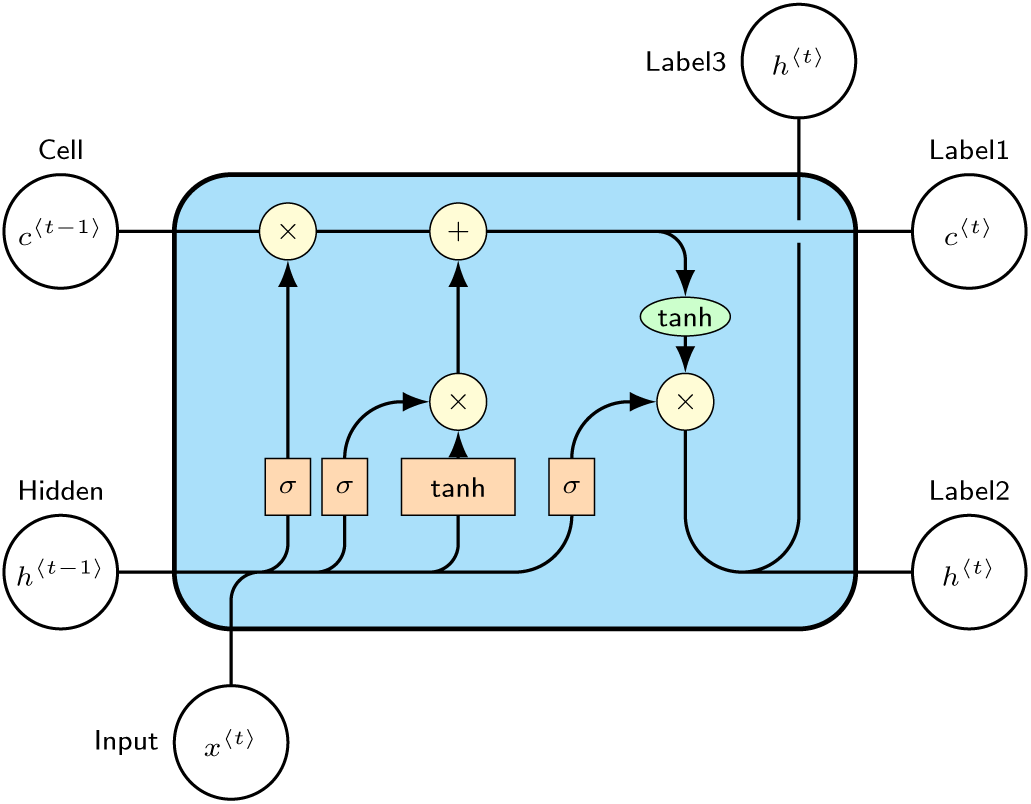
Diagram of Long Short-Term Memory (LSTM) cell architecture.

We used a standard LSTM architecture to ensure consistency across experiments and to isolate the impact of different input configurations (e.g., with or without climate and mobility covariates). The model consists of a single LSTM layer with 1000 units, followed by a dense output layer with one unit. It uses the Adam optimizer with ReLU activation, and is trained using the mean squared error (MSE) loss function for 2500 epochs. All input features were scaled using MinMaxScaler.

Given that hyperparameter selection is highly data-dependent, we did not conduct exhaustive tuning for each city. Our goal is to provide a generalizable and reproducible framework for dengue forecasting, rather than to optimize model performance for a specific location. In practice, public health analysts may further fine-tune the model architecture and training settings to suit local conditions.

### 2.5 Uncertainty estimation

Unlike statistical models that naturally provide confidence intervals (CIs) through parametric assumptions about the distribution of residuals, deep learning models such as LSTMs do not inherently generate confidence intervals, making it difficult to quantify the uncertainty in predictions. Understanding this uncertainty is crucial in epidemiological forecasting, as it provides public health authorities with risk assessments and informs decision-making for dengue outbreak preparedness.

To address this challenge, we implement an adaptive conformal prediction framework (Vovk, Gammerman, & Shafer, 2005; Balasubramanian, Ho, & Vovk, 2014), a robust, distribution-free statistical method that constructs prediction intervals with a predefined confidence level. Conformal prediction ensures that the true dengue case count falls within the estimated interval with a specified probability, providing valid uncertainty quantification regardless of the underlying model’s complexity or assumptions. This makes it particularly suitable for epidemiological time series forecasting, where traditional methods for constructing prediction intervals may not be applicable.

For each time *t* and forecast prediction horizon *h*, we define the nonconformity score as the residual between the actual dengue case count and the model’s predicted value:

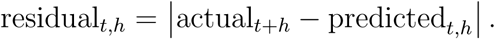

To estimate the distribution of residuals, we use a rolling window approach with window size *W*, capturing past prediction errors within a dynamically updated time frame. The set of residuals from time *t* − *W* to *t* for forecast horizon *h* is given by:

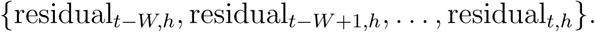

From these residuals, we compute quantiles to define the upper and lower prediction intervals. Specifically, for a given significance level *α* (e.g., 0.05 for a 95% uncertainty interval), the bounds are calculated as follows:

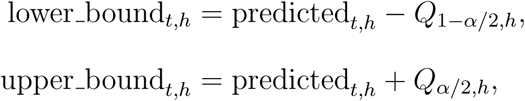

where *Q*_1−*α/*2*,h*_ and *Q_α/_*_2*,h*_ are the empirical quantiles of the residuals within the rolling window for the *h*-step ahead forecast. The quantiles are determined using:

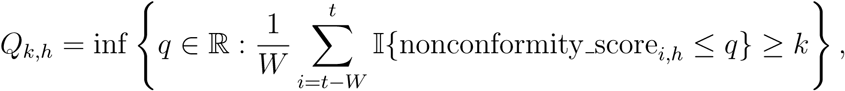

where Ι is the indicator function, ensuring that the estimated bounds maintain the desired coverage probability.

To enhance the reliability of the prediction intervals, we employ an adaptive calibration approach (Gibbs & Candes, 2021; Zaffran, Féron, Goude, Josse, & Dieuleveut, 2022). Specifically, we implement the Adaptive Generalized Conformal Inference (AgACI) framework (Zaffran et al., 2022), which adaptively calibrates upper and lower bounds using separate candidate sets. The dataset is split into a training set and a calibration set. The LSTM model is trained on the training set to generate initial forecasts, while the calibration set is used to compute nonconformity scores (residuals) from past predictions.

As new observations become available, the calibration set is dynamically updated, allowing prediction intervals to adapt to temporal variations in dengue transmission. This ensures that the conformal prediction intervals remain responsive to real-time epidemiological trends, reducing bias caused by shifting disease dynamics or changing climate conditions.

By integrating adaptive conformal prediction into our LSTM-based dengue forecasting model, we provide rigorous, model-agnostic uncertainty quantification, addressing a key limitation of deep learning-based time series models. To ensure interpretability for count-valued forecasts, we additionally apply a post-hoc truncation of negative lower bounds at zero, consistent with the non-negativity of dengue case counts. The resulting prediction intervals offer reliable confidence estimates, supporting more informed, risk-aware decision-making for dengue prevention and control strategies.

### 2.6 Model’s performance measures

To comprehensively assess the performance of the proposed model, we evaluate both dengue case forecasting accuracy and outbreak detection performance. The inclusion of both deterministic and probabilistic evaluation metrics ensures a robust assessment of the model’s predictive capability, providing valuable insights for public health decision-making.

#### 2.6.1 Forecasting accuracy of dengue cases

To quantify the accuracy of dengue case predictions, we employ three key metrics: Mean Absolute Error (MAE), Mean Absolute Percentage Error (MAPE), and Continuous Ranked Probability Score (CRPS). The MAE measures the average absolute deviation between predicted and actual dengue cases:

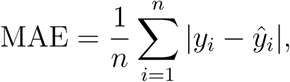

where *y_i_* represents the observed dengue cases, *y*^*_i_*represents the predicted values, and *n* is the number of observations. MAE provides an intuitive measure of forecast error magnitude, without considering directionality.

The MAPE expresses the error as a percentage, offering a relative assessment of prediction accuracy across different cases levels:

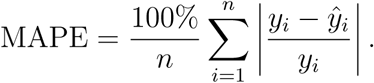

By normalizing errors relative to actual values, MAPE facilitates comparison across regions with varying dengue cases, making it particularly useful for large-scale forecasting evaluations.

To assess the reliability of the uncertainty-aware probabilistic forecasts, we use the Continuous Ranked Probability Score (CRPS), which evaluates the accuracy of the predicted probability distribution. CRPS is defined as:

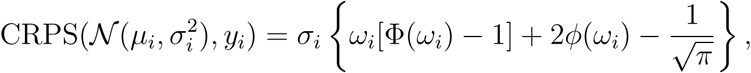

where *µ_i_* and *σ_i_* denote the predicted mean and standard deviation of dengue cases, respectively, and

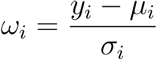

represents the normalized prediction error. Here, Φ(*ω_i_*) and *ϕ*(*ω_i_*) denote the cumulative distribution function (CDF) and probability density function (PDF) of the standard normal distribution, respectively. A lower CRPS value indicates well-calibrated forecasts with properly estimated uncertainty, ensuring that the predicted intervals accurately capture real dengue trends.

#### 2.6.2 Outbreak detection performance

In addition to evaluating numerical forecasting accuracy, we assess the model’s ability to predict dengue outbreaks by classifying each epidemiological week in 2023 as either an outbreak or a non-outbreak period. The outbreak classification performance is measured using Accuracy, Sensitivity (Recall), Specificity, and the F1 Score.

Accuracy, which measures the proportion of correctly classified outbreak and non-outbreak weeks, is given by

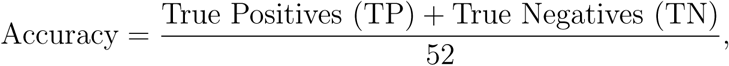

where 52 represents the total number of epidemiological weeks in 2023.

Sensitivity (Recall) quantifies the model’s ability to correctly detect actual out-breaks:

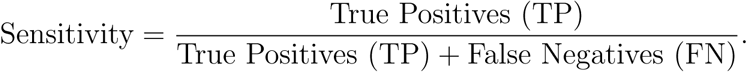

A high sensitivity ensures that the model effectively captures outbreak periods, minimizing the risk of failing to provide early warnings.

Specificity, on the other hand, evaluates how well the model avoids false alarms by correctly identifying non-outbreak weeks:

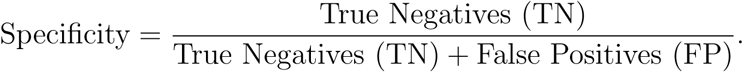

Ensuring high specificity is critical in preventing unnecessary public health interventions in weeks where no outbreak occurs.

Finally, the F1 Score, which balances precision and recall, is computed as

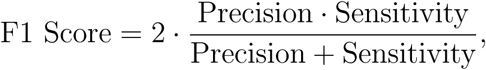

where precision is defined as

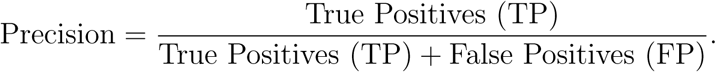

While sensitivity measures the ability to detect outbreaks and specificity reflects how well false alarms are avoided, the F1 Score provides a single metric that accounts for both, ensuring a balanced evaluation of the model’s outbreak detection performance. By integrating both numerical accuracy metrics for dengue case forecasting and classification metrics for outbreak detection, this evaluation framework ensures that the proposed model is both statistically sound and practically relevant for public health applications.

## 3 Results

In this section, we assess the performance of the proposed LSTM-based dengue forecasting model by evaluating its ability to predict both dengue case counts and outbreaks. To analyze the contribution of human mobility data, we compare our model against three baselines that incorporate different levels of information:

1. Baseline 1: LSTM using only historical dengue case data.
2. Baseline 2: LSTM incorporating both dengue case data and climate variables.
3. Baseline 3: LSTM integrating dengue case data, climate variables, and direct geographic neighborhood structure, which means incorporating lagged dengue case data from directly adjacent regions that share a geographic border.
4. Proposed Model: LSTM incorporating dengue case data, climate variables, and human mobility data.

By comparing these models, we aim to quantify the added value of mobility information in improving dengue prediction and outbreak detection. The results are presented in two parts: (1) forecasting performance for dengue case predictions and (2) outbreak prediction performance under both threshold-based outbreak definitions.

All analyses were implemented in Python, using TensorFlow 2.11 for model construction and training, along with standard libraries such as NumPy, Pandas, Matplotlib, and Scikit-learn for data processing and visualization.

### 3.1 Dengue case prediction performance

Table 2 presents the forecasting performance of the proposed model compared to the three baseline models across ten selected Brazilian cities. The evaluation metrics—Mean Absolute Error (MAE), Mean Absolute Percentage Error (MAPE), and Continuous Ranked Probability Score (CRPS)—are used to assess both the accuracy and reliability of the predictions.

**Table 2:** Comparison of forecasting performance across selected Brazilian cities using different models.

| City | State | Baseline 1 |  |  | Baseline 2 |  |  | Baseline 3 |  |  | Model |  |  |
| --- | --- | --- | --- | --- | --- | --- | --- | --- | --- | --- | --- | --- | --- |
|  |  | MAE | MAPE | CRPS | MAE | MAPE | CRPS | MAE | MAPE | CRPS | MAE | MAPE | CRPS |
| Manaus | AM | 98.16 | 42.89% | 22.74 | 80.38 | 31.32% | 16.72 | 78.31 | 30.00% | 16.58 | **75.45** | **29.95%** | **15.31** |
| Belém | PA | 28.11 | 40.09% | 6.00 | 24.77 | 32.30% | 5.20 | 25.64 | 30.03% | 5.19 | **23.98** | **29.28%** | **5.19** |
| Fortaleza | CE | 428.81 | 38.01% | 93.73 | 361.72 | 29.66% | 79.32 | 277.54 | 26.20% | 71.61 | **247.27** | **22.59%** | **68.95** |
| Salvador | BA | 386.72 | 39.24% | 79.06 | 299.63 | 30.95% | 56.97 | 252.09 | 25.14% | 52.54 | **228.55** | **23.95%** | **47.91** |
| Brasília | DF | 1480.98 | 35.05% | 327.45 | 1359.57 | 31.11% | 308.13 | 1276.06 | 27.24% | 281.61 | **1067.66** | **22.12%** | **213.89** |
| Goiânia | GO | 604.09 | 35.64% | 129.50 | 547.72 | 29.22% | 107.42 | 524.47 | 25.47% | 93.17 | **439.02** | **23.26%** | **88.87** |
| Belo horizonte | MG | 2360.34 | 39.28% | 480.91 | 1555.45 | 34.74% | 283.63 | 1501.73 | 28.24% | 267.42 | **1483.27** | **22.47%** | **257.55** |
| Rio de Janeiro | RJ | 1153.40 | 39.78% | 189.70 | 1210.43 | 31.60% | 170.82 | 1010.78 | 27.82% | 168.70 | **819.73** | **21.87%** | **158.93** |
| São Paulo | SP | 1812.24 | 35.65% | 348.72 | 1490.23 | 30.61% | 275.45 | 1230.09 | 24.84% | 227.07 | **1102.75** | **22.18%** | **205.74** |
| Curitiba | PR | 101.61 | 38.55% | 20.20 | 77.79 | 34.55% | 14.81 | 70.21 | 27.22% | 14.21 | **65.99** | **25.33%** | **13.11** |

Across all cities, Baseline 2 consistently outperforms Baseline 1, demonstrating that incorporating climate variables improves forecasting accuracy beyond relying on historical dengue case data alone. For example, in Manaus (AM), MAE decreases from 98.16 in Baseline 1 to 80.38 in Baseline 2, while MAPE drops from 42.89% to 31.32%, highlighting the influence of climatic factors in shaping dengue transmission patterns. Similarly, in Braśılia (DF), the inclusion of climate variables reduces the MAE from 1480.98 to 1359.57, and MAPE from 35.05% to 31.11%, confirming that climate information enhances temporal forecasting.

Further improvements are observed in Baseline 3, where adding geographic neighborhood effects leads to more accurate predictions across all cities. In Rio de Janeiro (RJ), MAE improves from 1210.43 (Baseline 2) to 1010.78 (Baseline 3), while CRPS decreases from 170.82 to 168.70, indicating that accounting for dengue spread in neighboring locations refines the forecasts. A similar trend is seen in Belo Horizonte (MG), where MAE is reduced from 1555.45 (Baseline 2) to 1501.73 (Baseline 3), and MAPE from 34.74% to 28.24%, showing that incorporating spatial transmission dependencies enhances prediction accuracy.

However, the proposed model consistently outperforms all baselines, demonstrating the added value of integrating human mobility data into dengue forecasting. The largest improvements are observed in highly connected urban centers, where mobility-driven transmission plays a significant role. In São Paulo (SP), the proposed model achieves an MAE of 1102.75, compared to 1230.09 in Baseline 3, and significantly reduces CRPS from 227.07 (Baseline 3) to 205.74, indicating improved uncertainty calibration. In Goiânia (GO), MAE improves from 524.47 (Baseline 3) to 439.02, and MAPE drops from 25.47% to 23.26%, further validating the role of mobility in disease forecasting.

Notably, even in cities with relatively lower absolute dengue case counts, such as Curitiba (PR), mobility information provides additional improvements. The proposed model achieves a MAPE of 25.33%, lower than the 27.22% in Baseline 3, while CRPS decreases from 14.21 to 13.11, reinforcing the importance of mobility effects even in regions with relatively lower transmission intensity.

Across all geographic regions, from the northern Amazonian cities (Manaus, Beĺem) to the southeastern megacities (Sāo Paulo, Rio de Janeiro) and southern urban centers (Curitiba), the proposed model consistently achieves the best performance. These improvements suggest that human mobility significantly contributes to dengue spread across connected regions, and its incorporation leads to more precise, reliable, and spatially-aware forecasting.

By explicitly modeling mobility-driven dengue importation, the proposed model achieves lower MAE and MAPE values, reflecting greater prediction accuracy, while simultaneously achieving lower CRPS values, indicating better-calibrated uncertainty estimates. These findings reinforce the critical role of human movement in dengue transmission dynamics and highlight the necessity of mobility-aware forecasting models for effective epidemic surveillance and early warning systems.

### 3.2 Outbreak detection performance

Tables 3 and 4 summarize the performance of all models in detecting dengue outbreaks, evaluated under the historical mean threshold and the mean plus two standard deviations threshold, respectively. The effectiveness of each model is measured using accuracy, sensitivity, specificity, and the F1 score.

**Table 3:** Comparison of dengue outbreak forecasting performance across selected Brazilian cities using different models under the historical mean threshold.

| City | State | Baseline-1 |  |  |  | Baseline-2 |  |  |  | Baseline-3 |  |  |  | Model Result |  |  |  |
| --- | --- | --- | --- | --- | --- | --- | --- | --- | --- | --- | --- | --- | --- | --- | --- | --- | --- |
|  |  | Accuracy | Sensitivity | Specificity | F1 Score | Accuracy | Sensitivity | Specificity | F1 Score | Accuracy | Sensitivity | Specificity | F1 Score | Accuracy | Sensitivity | Specificity | F1 Score |
| Manaus | AM | 0.8269 | 0.6842 | 0.9091 | 0.7429 | 0.9038 | 0.7222 | 1.0000 | 0.8387 | 0.8846 | 0.6842 | 1.0000 | 0.8125 | 0.9231 | 0.7647 | 1.0000 | 0.8667 |
| Belém | PA | 0.6731 | 0.0769 | 0.8718 | 0.1053 | 0.6923 | 0.0769 | 0.8974 | 0.1111 | 0.6731 | 0.0769 | 0.8718 | 0.1053 | 0.6923 | 0.0769 | 0.8974 | 0.1111 |
| Fortaleza | CE | 0.8654 | 0.0000 | 0.9375 | 0.0000 | 0.9231 | 0.5000 | 0.9583 | 0.5000 | 0.9231 | 0.6667 | 0.9388 | 0.5000 | 0.9615 | 0.7500 | 0.9792 | 0.7500 |
| Salvador | BA | 0.6346 | 0.6531 | 0.3333 | 0.7711 | 0.6538 | 0.6735 | 0.3333 | 0.7857 | 0.6346 | 0.6531 | 0.3333 | 0.7711 | 0.6923 | 0.7143 | 0.3333 | 0.8140 |
| Brasília | DF | 0.7500 | 0.6579 | 1.0000 | 0.7937 | 0.7692 | 0.6842 | 1.0000 | 0.8125 | 0.7308 | 0.6579 | 0.9286 | 0.7812 | 0.9423 | 0.9211 | 1.0000 | 0.9589 |
| Goiania | GO | 0.7500 | 0.4667 | 0.8649 | 0.5185 | 0.7885 | 0.6000 | 0.8649 | 0.6207 | 0.7500 | 0.4667 | 0.8649 | 0.5185 | 0.8846 | 0.6923 | 0.9487 | 0.7500 |
| Belo horizonte | MG | 0.7885 | 0.7317 | 1.0000 | 0.8451 | 0.7885 | 0.7317 | 1.0000 | 0.8451 | 0.7885 | 0.7317 | 1.0000 | 0.8451 | 0.7885 | 0.7317 | 1.0000 | 0.8451 |
| Rio de Janeiro | RJ | 0.6731 | 0.6731 | — | 0.8046 | 0.8645 | 0.8645 | — | 0.9278 | 0.8077 | 0.8077 | — | 0.8936 | 0.8846 | 0.8846 | — | 0.9388 |
| São Paulo | SP | 0.7308 | 0.7255 | 1.0000 | 0.8409 | 0.8846 | 0.8824 | 1.0000 | 0.9375 | 0.8462 | 0.8431 | 1.0000 | 0.9149 | 0.9038 | 0.9020 | 1.0000 | 0.9485 |
| Curitiba | PR | 0.8077 | 0.7368 | 1.0000 | 0.8485 | 0.8654 | 0.8158 | 1.0000 | 0.8986 | 0.8077 | 0.7368 | 1.0000 | 0.8485 | 0.9038 | 0.8484 | 1.0000 | 0.9296 |

**Table 4:** Comparison of dengue outbreak forecasting performance across selected Brazilian cities using different models under the mean plus two standard deviations threshold.

| City | State | Baseline-1 |  |  |  | Baseline-2 |  |  |  | Baseline-3 |  |  |  | Model Result |  |  |  |
| --- | --- | --- | --- | --- | --- | --- | --- | --- | --- | --- | --- | --- | --- | --- | --- | --- | --- |
|  |  | Accuracy | Sensitivity | Specificity | F1 Score | Accuracy | Sensitivity | Specificity | F1 Score | Accuracy | Sensitivity | Specificity | F1 Score | Accuracy | Sensitivity | Specificity | F1 Score |
| Manaus | AM | 0.7885 | 0.5000 | 0.9167 | 0.5926 | 0.8654 | 0.5625 | 1.0000 | 0.7200 | 0.8077 | 0.5000 | 0.9444 | 0.6154 | 0.8846 | 0.6000 | 1.0000 | 0.7500 |
| Belém | PA | 0.9231 | 0.0000 | 1.0000 | 0.0000 | 0.9231 | 0.0000 | 1.0000 | 0.0000 | 0.9231 | 0.0000 | 1.0000 | 0.0000 | 0.9615 | 0.0000 | 1.0000 | 0.0000 |
| Fortaleza | CE | 0.9423 | 0.0000 | 0.9800 | 0.0000 | 0.9615 | 0.0000 | 1.0000 | 0.0000 | 0.9615 | 0.0000 | 1.0000 | 0.0000 | 0.9615 | 0.0000 | 1.0000 | 0.0000 |
| Salvador | BA | 0.6538 | 0.6444 | 0.7143 | 0.7632 | 0.6731 | 0.6667 | 0.7143 | 0.7792 | 0.6538 | 0.6444 | 0.7143 | 0.7632 | 0.7308 | 0.7333 | 0.7143 | 0.8250 |
| Brasília | DF | 0.7692 | 0.6562 | 0.9500 | 0.7778 | 0.7692 | 0.6562 | 0.9500 | 0.7778 | 0.7692 | 0.6562 | 0.9500 | 0.7778 | 0.9808 | 1.0000 | 0.9500 | 0.9846 |
| Goiania | GO | 0.8462 | 0.2857 | 0.9333 | 0.3333 | 0.9038 | 0.5000 | 0.9565 | 0.5455 | 0.8462 | 0.2500 | 0.9545 | 0.3333 | 0.9231 | 0.5714 | 0.9778 | 0.6667 |
| Belo horizonte | MG | 0.7500 | 0.7105 | 0.8571 | 0.8060 | 0.7692 | 0.7179 | 0.9231 | 0.8235 | 0.7308 | 0.6842 | 0.8571 | 0.7879 | 0.7885 | 0.7179 | 1.0000 | 0.8358 |
| Rio de Janeiro | RJ | 0.8462 | 0.7879 | 0.9474 | 0.8667 | 0.9038 | 0.8485 | 1.0000 | 0.9180 | 0.8846 | 0.8182 | 1.0000 | 0.9000 | 0.9231 | 0.8788 | 1.0000 | 0.9355 |
| São Paulo | SP | 0.6731 | 0.6667 | 1.0000 | 0.8000 | 0.8462 | 0.8431 | 1.0000 | 0.9149 | 0.7885 | 0.7843 | 1.0000 | 0.8791 | 0.8846 | 0.8824 | 1.0000 | 0.9375 |
| Curitiba | PR | 0.7885 | 0.7297 | 0.9333 | 0.8308 | 0.7692 | 0.6944 | 0.9375 | 0.8065 | 0.7885 | 0.7429 | 0.8824 | 0.8254 | 0.8269 | 0.7778 | 0.9375 | 0.8615 |

Under the historical mean threshold (Table 3), the proposed model outperforms all three baselines across nearly all cities, achieving the highest accuracy, sensitivity, and F1 scores. This indicates that integrating human mobility data enhances outbreak detection by capturing imported cases that may contribute to local transmission surges. When comparing baselines, Baseline 2 generally performs better than Baseline 1, while Baseline 3 shows performance similar to Baseline 1 in most cases. This suggests that climate variables improve outbreak detection more effectively than geographic neighborhood effects. One possible reason is that neighboring areas may provide misleading signals about outbreak occurrence, as dengue transmission is not strictly confined to geographically adjacent regions but is instead influenced by human mobility patterns. As a result, relying solely on direct geographic neighbors can introduce incorrect outbreak classifications, whereas climate variables offer more consistent predictors of mosquito activity and disease spread.

For instance, in Braśılia (DF), the proposed model achieves an F1 score of 0.9589, significantly improving over Baseline 1 (0.7937) and Baseline 2 (0.8125). Similarly, in Sāo Paulo (SP), the proposed model attains the highest specificity (1.000) and an F1 score of 0.9485, demonstrating its reliability in distinguishing outbreak and non-outbreak periods.

Notably, in Rio de Janeiro (RJ), specificity cannot be calculated across models because, under this threshold, all weeks in 2023 are classified as outbreaks, meaning there are no true negative cases. This highlights that historical mean-based outbreak definitions may be less reliable in high-incidence regions, as they may overestimate outbreak occurrence in settings with persistently high dengue transmission.

Under the mean plus 2 standard deviations threshold (Table 4), all models exhibit a decline in performance, reflecting the increased stringency of the outbreak definition. With a higher threshold, fewer weeks are classified as outbreaks, making it more challenging for the models to correctly identify outbreak events.

Despite this, the proposed model maintains superior performance, achieving the highest F1 scores in most cities. The decline in sensitivity is particularly noticeable, as stricter thresholds result in fewer true positives, making it harder for the models to capture outbreak periods.

In the northern cities (e.g., Beĺem (PA) and Fortaleza (CE)), the sensitivity value drops to 0 across all models, indicating that no weeks in 2023 exceeded the outbreak threshold. This suggests that, under this stricter definition, these regions did not experience outbreaks based on past trends, reinforcing the importance of carefully selecting outbreak thresholds to avoid misclassification.

In cities with historically high dengue incidence, such as Rio de Janeiro (RJ) and São Paulo (SP), the proposed model still achieves high accuracy and specificity, with an F1 score of 0.9355 in Rio de Janeiro and 0.9375 in São Paulo, demonstrating its robustness even under more conservative outbreak definitions.

Across both outbreak thresholds, the proposed model consistently achieves the best balance between sensitivity and specificity, reinforcing the value of mobility-aware forecasting in detecting outbreaks more effectively than traditional climate-based or geographic proximity models. The results highlight that, while threshold selection significantly impacts outbreak classification, incorporating human mobility information enhances the model’s ability to identify epidemiologically relevant transmission patterns, particularly in highly connected urban centers.

## 4 Discussion

This study addresses the challenges of dengue forecasting by developing an LSTM-based predictive model that integrates human mobility data, climate variables, and historical dengue cases to enhance prediction accuracy. Unlike traditional approaches that rely solely on epidemiological trends or static spatial structures, our model explicitly incorporates imported dengue cases via mobility-adjusted transmission dynamics, allowing for a more realistic representation of disease spread. The comparative analysis with baseline models demonstrates the effectiveness of integrating human mobility, as reflected in the improvements in both dengue case forecasting accuracy and out-break prediction performance. These findings highlight the importance of capturing mobility-driven transmission, particularly in densely connected urban regions where human movement significantly influences dengue dynamics. Although our mobility-adjusted variable is not intended to establish causality, the observed improvements in model performance support its utility as a predictive feature for capturing inter-city transmission risk.

Although this study focuses on only 10 selected cities, the proposed framework is highly generalizable and can be extended to other cities, regions, and even different countries. The core methodology—incorporating mobility-driven transmission risks into deep learning-based forecasting—is not geographically constrained and can be adapted to any location where reliable mobility, climate, and epidemiological data are available. This adaptability makes the model a promising tool for broader disease surveillance and outbreak forecasting efforts beyond dengue, including other vector-borne diseases such as Zika and Chikungunya, which share similar transmission pathways. Future studies could expand the model to nationwide or global scales by integrating cross-border travel data, heterogeneous mobility sources (e.g., mobile phone data), and regional variations in dengue risk factors.

Despite its advancements, this study has several limitations that warrant further exploration. One potential limitation lies in the reliability of dengue case reports, which may be affected by underreporting and reporting delays. However, since this study focuses on city-level data from ten major urban centers, where surveillance systems are relatively robust, the impact of data inconsistencies is expected to be lower. Nevertheless, if applied to smaller municipalities or regions with weaker surveillance infrastructure, the model’s performance could be influenced by systematic biases in dengue reporting. Future studies expanding to such regions may consider data augmentation techniques or syndromic surveillance integration (e.g., online search trends, pharmacy sales data, or social media signals) to mitigate potential reporting gaps (Höhle & an der Heiden, 2014; Rotejanaprasert, Ekapirat, Areechokchai, & Maude, 2020; Xiao, Soares, Bastos, Izbicki, & Moraga, 2024).

Another limitation stems from the granularity and completeness of human mobility data. While the current study incorporates intercity travel flows, finer-scale mobility patterns within cities—such as commuting behavior, residential mobility, or seasonal travel trends—are not explicitly captured. Since dengue transmission often occurs at neighborhood or household scales, short-distance mobility plays a crucial role in sustaining local transmission cycles. Future studies could integrate mobile phone-based mobility datasets, transportation network data, or anonymized GPS trajectories to refine estimates of intra-city human movement and its contribution to localized dengue transmission (Ihantamalala et al., 2018; Massaro, Kondor, & Ratti, 2019). Additionally, our approach currently treats all mobility routes equally, but travel behaviors may vary seasonally or socioeconomically, influencing transmission in complex ways. Adapting time-varying mobility weights based on external mobility trends (e.g., holidays, migration events, or travel restrictions (Islam & Hu, 2024)) could further improve forecasting accuracy.

A third limitation relates to the assumption of homogeneous vector distribution and transmission efficiency across different regions. While the model accounts for imported cases in proportion to origin-city prevalence and population, it does not explicitly consider local mosquito abundance, environmental suitability, or vector control interventions that could influence whether imported cases lead to sustained out-breaks (Organization et al., 2012). For instance, an area with high human mobility but low mosquito density may not experience the same outbreak risks as a high-mobility, high-vector-density region. Future research could incorporate vector surveillance data—such as mosquito density indices (Bowman, Runge-Ranzinger, & McCall, 2014), insecticide resistance data, or spatially explicit climate suitability models—to refine risk estimates based on both human-driven and vector-driven transmission factors.

In our study, we only used temperature and humidity as climate predictors, as these were the variables consistently available from the InfoDengue platform. This choice is also consistent with our previous national-scale research (Chen & Moraga, 2025b), where temperature and humidity emerged as the most important predictors across regions. However, if conditions permit, future studies could incorporate additional external climate variables—such as precipitation, thermal range, or vegetation indices—from high-resolution reanalysis or satellite-based datasets to further enhance model performance.

Additionally, we included the Continuous Ranked Probability Score (CRPS) as a proper scoring rule to assess probabilistic forecast performance by assuming a continuous distribution which may not fully capture the discrete, non-negative nature of dengue case counts. Future work could adopt the Weighted Interval Score (WIS), which aligns better with quantile-based prediction intervals such as those generated through conformal prediction (Bracher, Ray, Gneiting, & Reich, 2021; Coĺon-González et al., 2021).

By addressing these limitations, future research can refine the model’s ability to capture both spatial and temporal complexities of dengue transmission, ultimately enhancing its applicability for real-time epidemic forecasting and public health decision-making.

## 5 Conclusion

This study presents a comprehensive and scalable framework for dengue forecasting that integrates human mobility data, climate variables, and historical dengue cases into an LSTM-based predictive model. By explicitly incorporating imported dengue cases through mobility-adjusted transmission dynamics, the proposed approach improves upon traditional forecasting methods that rely solely on epidemiological time series or static spatial structures. The results demonstrate that incorporating human movement patterns enhances both dengue case prediction accuracy and outbreak detection performance, underscoring the importance of mobility-driven disease transmission modeling.

While tested on ten major cities, the approach is generalizable to other regions and diseases influenced by human movement and climate factors. Future work will focus on enhancing mobility granularity, integrating vector surveillance data, and expanding to broader geographic scales.

By advancing data-driven and mobility-aware epidemic forecasting, this study contributes to the development of more effective dengue early warning systems, supporting proactive public health interventions and targeted outbreak response strategies.

## Declarations

### CRediT authorship contribution statement

**Xiang Chen**: Conceptualization, Methodology, Software, Formal Analysis, Investigation, Data Curation, Writing – Original Draft, Writing – Review & Editing, Visualization. **Paula Moraga**: Supervision, Conceptualization, Methodology, Writing – Review & Editing, Project Administration, Funding Acquisition.

### Ethical Approval and Consent to participate

Not applicable.

### Consent for publication

Not applicable.

## Data Availability

All data used in this study is open access and freely available on the internet, see the Methodology section for details.

## Acknowledgements

Not applicable.

